# Effects of a diverse prebiotic fibre blend on inflammation, the gut microbiota, and affective symptoms: A pilot open label randomised controlled trial

**DOI:** 10.1101/2024.02.12.24302681

**Authors:** C.V. Hall, P Hepsomali, B Dalile, L Scapozza, T. Gurry

## Abstract

Emerging evidence suggests that low-grade systemic inflammation plays a key role in altering brain activity, behaviour, and affect. Modulation of the gut microbiota using prebiotic fibre offers a potential therapeutic tool to regulate inflammation, mediated via the production of short-chain fatty acids (SCFAs). However, the impact of prebiotic consumption on affective symptoms, and the possible contribution from inflammation, gut symptoms, and the gut microbiome, is currently underexamined. In this 12-week study, the effects of a diverse prebiotic blend on inflammation, gut microbiota profiles, and affective symptoms in a population with Metabolic Syndrome (MetS) was examined. Sixty patients meeting the criteria for MetS were randomised into a treatment group (n = 40), receiving 10g per day of a diverse prebiotic blend and healthy eating advice and a control group (n = 20), receiving healthy eating advice only. Our results showed a significant reduction in C-reactive protein (CRP), alongside improvements in self-reported affective scores in the treatment compared to the control group. While there were no differences in relative abundance between groups at week 12, there was a significant increase from baseline to week 12 in *Bifidobacterium* and *Parabacteroides* in the treatment group, both of which are recognised as SCFA producers. Multivariate regression analyses further revealed that changes in affective scores were positively associated with both gastrointestinal symptoms and CRP. Together, this study provides preliminary support for the use of a diverse prebiotic blend for mood, stress, and anxiety.

## Introduction

Metabolic syndrome (MetS) is defined as a cluster of risk factors including visceral or central obesity, glucose dysregulation, hypertension, and dyslipidaemia^(1)^. Alongside metabolic abnormalities, MetS is strongly associated with low-grade systemic inflammation, which is thought to play a critical role in the development of mood, stress, anxiety, and sleep disorders and/or symptomology^(2-5)^. As such, there is a strong bidirectional relationship between MetS, with anxiety disorders^(4)^ and major depression^(6)^. As part of this underlying mechanism, pro-inflammatory cytokines communicate with the central nervous system (CNS) by activating receptors on vagal afferents or by generating intermediates at the blood–brain interface (BBB)^(7)^. The brain recognises and interprets inflammation as a signal of illness, which collectively gives rise to “sickness behaviours”, including social avoidance, anhedonia, fatigue, anxiety, and depressed mood^(8-10)^. While sickness behaviours were initially studied in the context of acute inflammation (i.e., infection), more recent work shows that repeated or long-term exposure to low-grade systemic inflammation, as seen in conditions like MetS, is associated with an increased risk of future depressive and anxiety symptoms^(10-13)^. There is, therefore, a growing interest to develop novel, simple, and cost-effective interventions that can target systemic inflammation to improve long-term psychological health.

Recent work suggests that the gut microbiota and its metabolites can have a significant effect on attenuating low-grade systemic inflammation^(14)^, offering a potential therapeutic avenue to improve affective symptoms in MetS. One of the primary mechanisms by which the gut microbiota influences inflammation is via the production of short-chain fatty acids (SCFAs) during the microbial fermentation of prebiotic fibre^(15)^. SCFAs interact with virtually all pathways mediating gut-brain communication, including neuronal (vagal nerve), humoral, endocrine, and immune pathways^(16)^. Specifically, SCFAs can affect the CNS by regulating inflammation both locally^(17)^ and at the blood-brain interface (BBB)^(7, 13)^, rapidly signalling to the brain via enteroendocrine-mediated vagal signalling^(18, 19)^, or affecting gene expression by acting as histone deacetylase inhibitors (HDACs)^(20)^. To leverage the proposed benefits of SCFAs, prebiotic supplements have been utilised to investigate effects on mood, anxiety, stress, and cognition^(21-27)^. Importantly, these studies have been performed in sub-clinical or healthy populations, suggesting that the benefits of prebiotic interventions extend beyond major neuropsychiatric conditions. While early findings are promising, they often fail to assess broader gastrointestinal (GI), inflammatory, and microbial changes that may parse psychological improvements. In this study, we assessed the effects of a diverse prebiotic blend on systemic inflammation, GI symptoms, self-reported affective scores, and the gut microbiota, in a population with MetS.

## Methods

### Participants

In total, 60 participants (53.9 ± 9.8 years [mean ± SD]; 62% female) who met the International Diabetes Federation (IDF) criteria for MetS^(1)^ and passed the screening and eligibility assessments were randomised to either the treatment (*n* = 40) or control (*n* = 20) group, stratified by age and sex. Detailed inclusion and exclusion criteria are reported in **Supplementary Note 1**.

### Ethical statement

The study was approved by the NHS Human Research Ethics Committee of City and East (reference: 23/LO/0515). The study was registered on ClinicalTrials.gov (registration number: NCT06216626). Written informed consent was obtained for all participants in accordance with the Declaration of Helsinki.

### Study Design

The study (conducted in the United Kingdom between 19 June 2023 to 22 December 2023) was a 12-week open-label parallel randomised controlled trial (RCT), with two groups: a control group who received healthy eating advice and a treatment group who received healthy eating advice and consumed a prebiotic fibre supplement (10g/day) (**Supplementary Note 2 and Table 1**). The study was a decentralised design, where participants completed all study requirements via video call and self-administered sample collection kits. Study requirements were completed during a pre-intervention baseline session (baseline), and a follow-up session at the end of the 12-week intervention (week 12). All participants had a video call with a member of the research team prior to the baseline session to confirm they understood the instructions for completing questionnaires, sample collection, and daily prebiotic intake.

**Table 1.** Baseline participant demographics.

| Demographics | Treatment (n = 37) | Control (n = 16) | p-value |
| --- | --- | --- | --- |
| Gender | 27 female (67.5%) | 9 female (56%) | 0.38<sup>a</sup><br>($X^2 = 0.77$) |
| Age (years) | 53.6 ± 10.1 | 53.9 ± 9.3 | 0.90<sup>b</sup> |
| BMI (kg/m<sup>2</sup>) | 31.5 ± 9.6 | 29.6 ± 4.1 | 0.38<sup>b</sup> |
| Waist circumference (cm) | 103.0 ± 15.0 | 97.8 ± 7.2 | 0.24<sup>b</sup> |
| Hip circumference (cm) | 113.7 ± 13.9 | 105.4 ± 11.1 | **0.05<sup>b</sup>** |
| Current smokers (n) | 4 (10.8%) | 2 (12.5%) | 1.0<sup>a</sup><br>($X^2 = 6.7 \times 10^{-32}$) |
| Pre-diabetes diagnosis (n) | 13 (35.1%) | 8 (50%) | 0.48<sup>a</sup><br>($X^2 = 0.50$) |
| Daily fibre intake (g) | 17.2 ± 6.9 | 15.0 ± 4.8 | 0.29<sup>b</sup> |
| GAD-7 | 6.7 ± 5.4 | 8.3 ± 5.6 | 0.36<sup>b</sup> |
| PHQ-9 | 7.5 ± 6.1 | 9.3 ± 7.0 | 0.37<sup>b</sup> |

Mean ± SD; total number (percentage); <sup>a</sup> Pearson's Chi-squared test; <sup>b</sup> Parametric t-test. Body mass index (BMI); Generalised Anxiety Disorder 7-item scale (GAD-7); Patient Health Questionnaire-9 (PHQ-9).

### Prebiotic treatment

The prebiotic blend was powdered, unflavoured, and given to the participants in 300g packets lasting for 30 days each (total 3 packets per participant). A 10g scoop was included in each packet and participants were advised to consume one level scoop at any time of the day. Participants were provided with examples on how to consume the prebiotic blend (e.g., breakfast cereal, coffee, tea, water). To minimise participant withdrawal and ensure consistent use, they received a weekly survey via email, asking them to confirm that they had taken the prebiotic blend on each day of the week. If they failed to consume the prebiotic on any given day, they were prompted to provide a reason. This prebiotic blend has not been studied formally in a cohort of participants with MetS. Therefore, a 2:1 treatment allocation was adopted to yield more information about potential tolerance and side effects in MetS, while controlling cohort size for practical purposes.

### Healthy eating advice

All participants were provided with healthy eating advice prior to starting the 12-week intervention. Dietary recommendations were available via the Thriva online portal and were consistent with the Heart UK’s Healthy Eating Guidelines for MetS (**Supplementary Note 2**). These recommendations emphasised a Mediterranean diet, rich in fruit, vegetables, and healthy fats (omega-3 fatty acids), while reducing refined sugar, salt, processed foods, and alcohol intake. The Mediterranean diet is considered gold-standard dietary approach for participants with MetS and/or poor mental health^(28, 29)^.

### Questionnaires

Participants completed previously validated questionnaires including the 18-item FiberScreen^(30)^, Gastrointestinal Symptom Rating Scale (GSRS)^(31)^, Perceived Stress Scale (PSS)^(32)^, Depression Anxiety and Stress Scale (42-item) (DASS)^(33)^, Patient Health Questionnaire-9 (PHQ-9)^(34)^, and Generalised Anxiety Disorder (7-item) (GAD-7)^(35)^ at both timepoints.

To assess the effect of the intervention on affective scores, we assessed changes in the DASS and PSS scales. The DASS is a 42-item self-report instrument that is based on a dimensional, rather than categorical, assessment of psychological symptoms, providing high inter-subject variability in non-clinical and sub-clinical populations^(36)^. Critically, the DASS can clearly distinguish between the three negative affective states of depression, anxiety, and stress^(37)^. The PSS is a 10-item self-report instrument which asks participants how different situations affect feelings and perceived stress in the previous month^(32)^. This scale has also been validated in sub-clinical populations and also provides high inter-individual variability^(38)^.

The PHQ-9 and GAD-7 are clinical tools that are used for screening, diagnosing, and monitoring depression and generalised anxiety disorders, respectively. In this study, these assessments were performed to (a) rule out severe neuropsychiatric disorders at baseline, and (b) assess the development of major neuropsychiatric disorders during the intervention which may require withdrawal from the study and/or ongoing monitoring from the participant’s primary care team.

### Blood sample collection and processing

Finger-prick capillary blood sampling kits (Thriva Limited, UK) were shipped to the participant’s chosen address, with written instructions on correct usage. Blood samples (600uL) were collected after an 8-hour minimum fast. Participants were advised to return their sample immediately after collection using a prepaid envelope. Once samples arrived at the lab, Serum Separator Tubes (SST) were centrifuged to separate the blood serum within the sample for testing. This occurs immediately after arriving at the lab to prevent sample decay. The samples are then processed within a 48-hour window. CRP and lipid profiles were tested via a Roche Cobas c503 platform. Previous work has shown high intraclass correlation coefficients between self-sampling of capillary blood compared to venous analysis, as well as high patient tolerability^(39, 40)^.

### Stool sample collection and 16S sequencing

Participants were provided with a stool sampling kit (Carbiotix AB (publ), Lund, Sweden), including a sample collection tube, sample swab, and instruction card, to use at home. Participants were instructed to collect the sample within 24 hours of completing other study requirements (blood sample and questionnaires) at baseline and week 12. Each stool sample was labelled and stored in a −80°C freezer until sample processing. Stool extraction was performed by homogenisation of sample (3000 rpm, 2 minutes) prior to the removal of 200uL of sample to be subjected to NA extraction on the kingfisher flex 96 system. Primers targeting the V4 region of the 16S gene were used. 16S rRNA sequencing was performed on an Illumina MiSeq platform^(41, 42)^.

### 16S data processing and analysis

Demultiplexed FASTQ files were processed using QIIME2 2020.2 (https://qiime2.org)^(43)^. Reads were quality filtered with a cut-off quality score of 20 and trimmed to a length of 150. Operational Taxonomic Units (OTUs) were generated by denoising with Deblur^(44)^. For taxonomic structure analysis, taxonomy was assigned to OTUs using a pre-trained Naïve Bayes classifier and the q2-feature-classifier plugin against the GreenGenes (gg_13_5) 16S rRNA gene sequencing database^(45)^. Samples were rarefied to a read depth of 10,000 for diversity analyses. Wilcoxon signed-rank tests was used to test for group differences in Shannon diversity and Chao1 richness measures. Beta-diversity, assessed using Bray Curtis distance, was used to compare between-group and within-group differences using Permutational Multivariate Analysis of Variance test (ADONIS2). Microbiome Multivariable Associations with Linear Models 2 (MaAsLin2)^(46)^ was used to assess (A) the changes in microbial abundance (collapsed at genus level) from baseline to week 12 in the treatment arm; and (B) the changes in microbial abundance (collapsed at genus level) between the treatment and control arm at week 12. Covariates including sex, age, and BMI, were included as fixed effects, and participant ID was included as a random effect (for model A only). Features were included if they had at least 10% non-zero values (across samples) and a minimum relative abundance threshold of 0.0001, both validated parameter settings in MaAsLin2. Significant features with *p* < 0.05 and *q* < 0.25 were considered statistically significant.

### Primary and secondary endpoints

The primary endpoint was the change in CRP from baseline to week 12. Key secondary endpoints included changes in DASS-S, DASS-D, DASS-A, PSS, gut microbiome profiles, lipid profiles, and systolic and diastolic blood pressure from baseline to week 12.

### Tolerability and safety

Participants were asked to record whether they experienced any side-effects or medical events since starting the intervention, recorded via online surveys at the end of weeks 4, 8, and 12. If they answered yes, Severe Adverse Events (SAE) and Adverse Events (AE) were documented, reported, and reviewed for relatedness and expectedness within 24 hours.

### Statistical tests

Data were analysed using R software (V4.3.1). Statistical analyses were performed based on an intention-to-treat analysis. Group differences in baseline characteristics were assessed using χ2-tests for categorical variables, and parametric t-tests or Wilcoxon signed-rank tests for quantitative variables. For all comparisons, we tested the normality and homoscedasticity. For the primary and secondary endpoints, an unpaired two-tailed t-test was used to compare the change from baseline to week 12 (i.e., week 12 – baseline) between the treatment and control groups. The choice to use unpaired t-tests was adopted to mitigate the influence of baseline differences and to ensure robustness to unequal sample sizes. Stepwise multiple linear regression models were performed to estimate the association between changes in gastrointestinal symptoms and CRP with changes in affective symptoms. The variable CRP was square root transformed prior to analyses. The models combined both groups (control and treatment) and included a regressor which adjusted for the effects of repeated measures. To confirm that multiple regression results remained consistent, we performed a sensitivity analysis which adjusted for the effects age, sex, and BMI (**Supplementary Table 2)**. A p-value < 0.05 was considered statistically significant. As a pilot study, our sample size was informed based on previous work assessing changes in our primary outcome, CRP, following a prebiotic or dietary intervention in a similar population. Results from previous studies in MetS (*n* = 52^(47)^; *n* = 50^(48)^), pre-diabetes (*n* = 51^(49)^), and non-alcoholic fatty liver (*n* = 66^(50)^) suggest that >50 participants is an adequate sample size to capture meaningful differences in our outcome measures. For feasibility purposes, a sample of 60 participants with MetS, with overrepresentation in the treatment relative to control group (2:1), was chosen.

## Results

### Baseline patient characteristics

Participant demographics in the treatment and control groups are reported in **Table 1**. Groups were matched except for hip circumference, which was not a matched variable for inclusion or randomisation (**Table 1**). A total of 53 participants (37 treatment; 16 control) completed the baseline and week 12 assessments. Reasons for participant dropouts included: lost to follow up (2 treatment; 2 control) and withdrawal with no reason provided prior to starting the intervention (1 treatment; 2 control).

### Adherence, tolerability, and safety

On average, participants in the treatment group consumed the prebiotic blend 73.0 ± 28.5% (mean ± SD) of the total intervention time. The treatment was well tolerated, with a larger reduction in total GSRS scores reported in the treatment (-0.34 ± 0.61) compared to the control (-0.06 ± 0.41), although not reaching statistical significance (*t_31.37_* = -1.84, *P* = 0.08, *d* = 0.50). When looking at the subscales of the GSRS, we only observed a significant reduction in abdominal pain in the treatment group (-0.41 ± 0.63) compared to the control (0.23 ± 0.53) (*t_24.59_* = -3.52, *P* = 0.002, *d* = 0.30) (**Supplementary Table 3**). There were no SAEs related to the treatment, and no participants withdrawing from the study due to product-related side-effects or disliking the taste. AEs reported were consistent with those typically seen in prebiotic-based interventions, including bloating, gas, and constipation (**Supplementary Table 4**).

### Dietary fibre intake

While dietary advice provided to participants emphasised the importance of a plant-based diet, we observed no significant change in dietary fibre intake (grams) from baseline to week 12 between the treatment (0.82 ± 5.57) and control (-0.33 ± 5.57) groups (*t_21.07_* = -0.64, *P* = 0.53, *d* = 0.21) (excluding the prebiotic blend for the treatment group).

### Primary endpoint

The change in CRP (mmol/L) from baseline to week 12 was significantly different between the treatment (-0.58 [-9.96 to 2.63]) and control groups (0.37 [-3.64 to 3.32]) (*W* = 98, *P* = 0.03, *HLE* = 0.43) (**Fig 1**).

**Fig 1.**
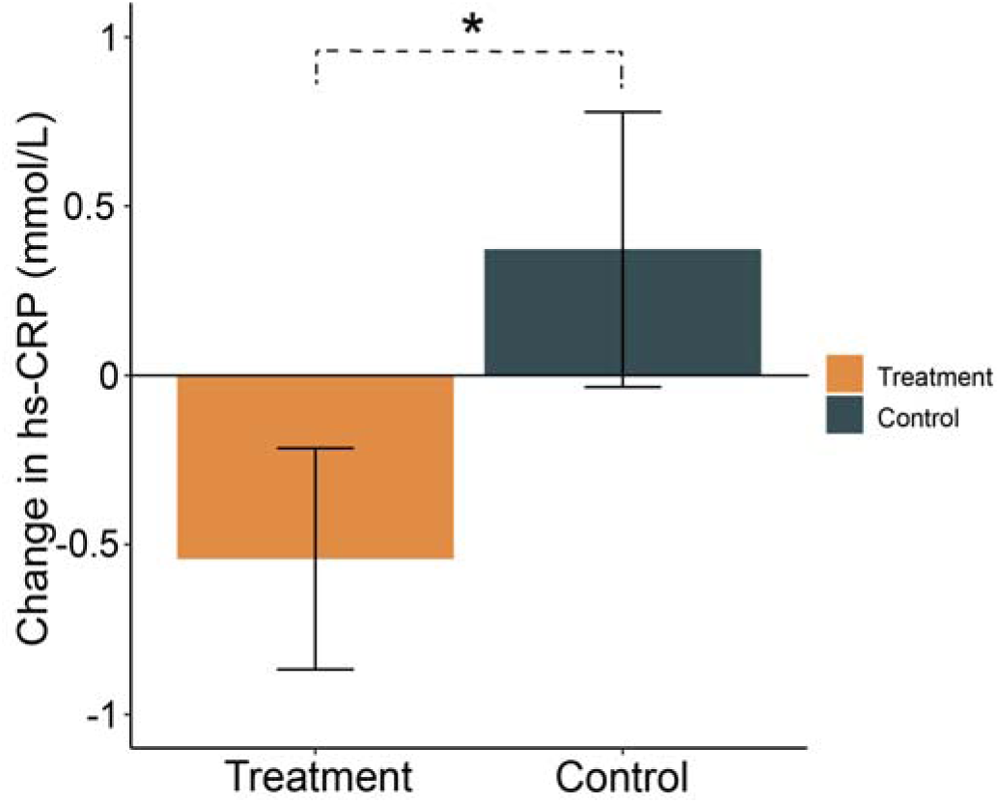
Changes from baseline to week 12 in C-reactive protein (CRP) (mmol/L) in the treatment (orange) and control (green) groups. Mean and standard errors are shown. * indicates *p*-value < 0.05.

### Affective scores

There was a significant difference between groups from baseline to week 12 in PSS (*t_29.03_* = - 3.15, *P* = 0.004, *d* = 0.88), DASS-S (*t_29.13_* = -2.81, *P* = 0.01, *d* = 0.76), DASS-A (*t_38.4_* = -2.31, *P* = 0.03, *d* = 0.57), and DASS-D (*t_43.64_* = -2.10, *P* = 0.05, *d* = 0.46) (**Fig. 2a-d**; **Supplementary Table 2**).

**Fig 2.**
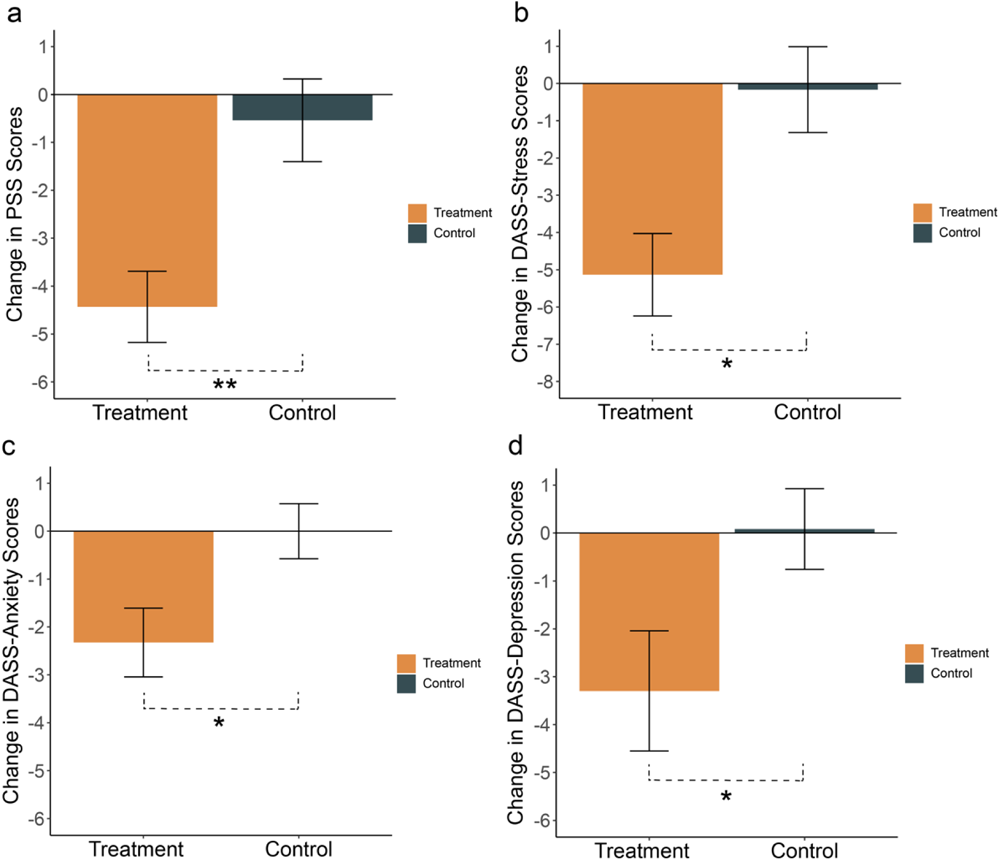
Changes from baseline to week 12 in (a) Perceived Stress Scale (PSS), (b) Depression, Anxiety, and Stress Scale 42-item (DASS) - Stress (DASS-S), (c) DASS - Anxiety (DASS-A), and (d) DASS-Depression (DASS-D) scores in the treatment (orange) and control (green) groups. Mean and standard errors are shown. * indicates *p*-value < 0.05 and ** indicates *p*-value < 0.005.

### Gut microbiome profiles

We observed no significant change from baseline to week 12 between groups in Shannon diversity (*W* = 264, *P* = 0.46, *HLE* = 0.06) or richness (*W* = 232, *P* = 0.17, *HLE* = 3.5). We also found no significant main or interaction effects in beta diversity (Bray Curtis) (*F* = 1.65, *P* = 0.08, *perms* = 9999). Multivariate analyses using MaAsLin2 revealed significant increases in *Bifidobacterium* (*p_FDR_* = 0.007, *q* = 0.24) and *Parabacteroides* (*p_FDR_*= 8.79 x 10^-5^, *q* = 0.003) in the treatment group following the prebiotic intervention, but no significant differences between groups at week 12.

### Blood pressure and lipid profiles

We observed a significant reduction in systolic BP from baseline to week 12 in the treatment group compared to control (*t_40.67_* = -2.21, *P* = 0.03, *d* = 0.29). No significant changes were observed between groups for diastolic BP (*t_43.65_* = 0.27, *P* = 0.79, *d* = 0.20) total cholesterol (*t_14.85_* = 0.37, *P* = 0.72, *d* = 0.75), triglycerides (*t_18.34_* = 1.71, *P* = 0.11, *d* = 0.15), LDL-C (*t_14.86_* = 0.44, *P* = 0.67, *d* = 0.18), or HDL-C (*t_24.91_* = -0.51, *P* = 0.62, *d* = 0.12) (**Supplementary Table 2**).

### Relationship between gastrointestinal symptoms, CRP, and affective symptoms

Results from our forward stepwise linear regression analyses showed significant multivariate associations between gut and inflammatory markers with higher scores on all six psychological scales (**Table 2**). Specifically, across all six models we found that changes in stress, anxiety, and depressive scores could be explained by changes in CRP and one or more gastrointestinal symptoms (**Fig. 3** & **Table 2**).

**Fig 3.**
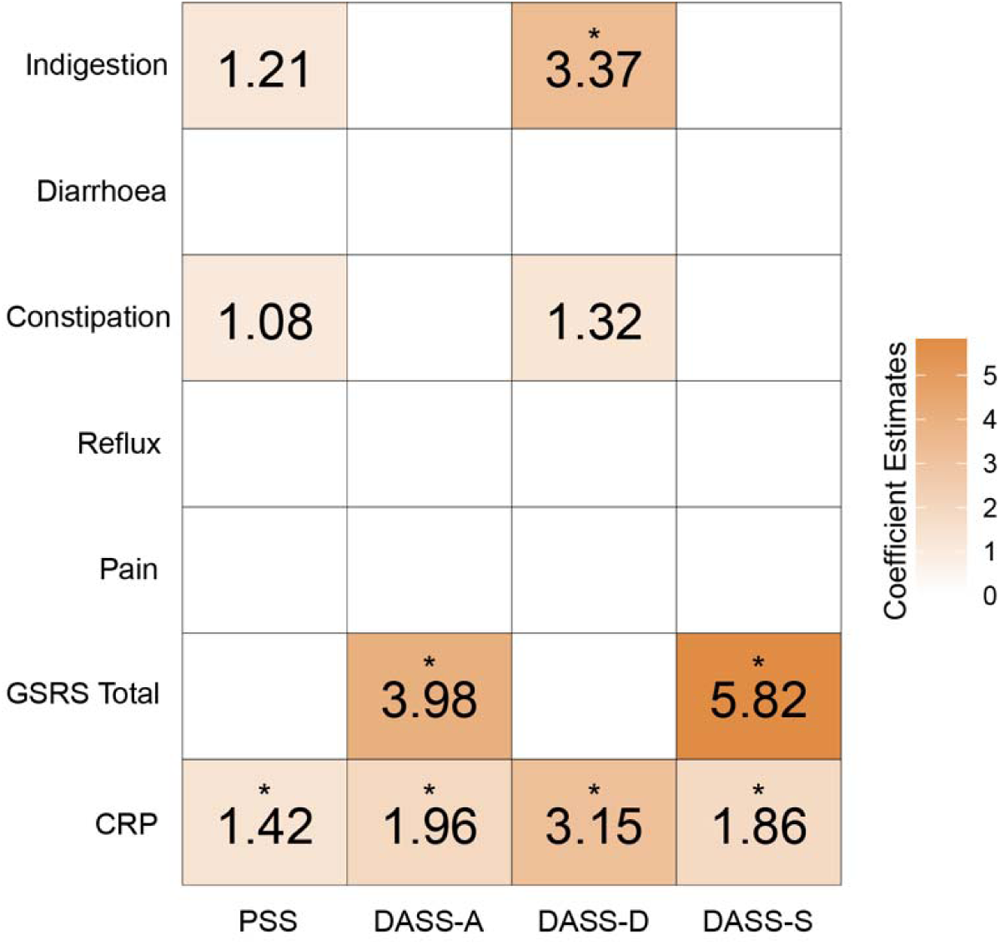
Heatmap showing multiple regression coefficient estimates representing the association between affective scores, with gastrointestinal and inflammation measures. Each column represents a different multiple regression model. Perceived Stress Scale (PSS); Depression, Anxiety, and Stress Scale 42-item - Anxiety (DASS-A); DASS-Depression (DASS-D); DASS-Stress (DASS-S); Gastrointestinal Symptom Rating Scale (GSRS); C-Reactive Protein (CRP). * indicates *p* < 0.05 (Bonferroni corrected).

**Table 2.** Results from multiple regression analyses.

| **Model** | **F-statistic** | **R<sup>2</sup>** | **Adjusted R<sup>2</sup>** | **p-values** |
| --- | --- | --- | --- | --- |
| PSS | 6.09 | 0.22 | 0.18 | **0.001** |
| DASS-A | 28.14 | 0.39 | 0.38 | **2.13 x 10<sup>-9</sup>** |
| DASS-D | 16.97 | 0.37 | 0.35 | **5.52 x 10<sup>-8</sup>** |
| DASS-S | 12.01 | 0.21 | 0.20 | **0.0001** |

Perceived Stress Scale (PSS); Depression, Anxiety, and Stress Scale 42-item (DASS)-Stress (DASS-S); DASS-Anxiety (DASS-A); DASS-Depression (DASS-D). All p-values are Bonferroni-corrected.

## Discussion

Our study showed significant changes in inflammation, the gut microbiome, and affective symptoms following a 12-week prebiotic fibre intervention with healthy diet advice in MetS, relative to participating receiving dietary advice only. Our primary outcome, CRP, exhibited a significant reduction from baseline to week 12 in the treatment group relative to the control. This reduction is consistent with two independent meta-analyses in overweight and obesity, showing reductions in circulating CRP following a fibre-rich food intervention or supplement^(51, 52)^. Another meta-analysis combining participants with underlying inflammatory conditions, including type 2 diabetes, hypercholesterolemia, liver disease, inflammatory bowel diseases, and obesity, showed similar decreases in CRP following prebiotic oligosaccharides compared with control^(53)^. Recent work by our group observed larger reductions in CRP when using a similar combination of prebiotic fibres in pre-diabetes^(49)^. The discrepancy between this work and our current study could, in part, be explained by a larger daily dose (20g), a longer intervention duration (16 weeks), or higher baseline CRP levels.

We observed significant improvements in the PSS, DASS-S, DASS-D, and DASS-A in the treatment compared to the control. While our affective outcomes were based on sub-clinical (i.e., no diagnosed neuropsychiatric disorders) scales, we also observed that the GAD and PHQ scales showed reductions in the treatment group too. Specifically, the treatment group shifted from the mild to minimal anxiety bracket (GAD), and from the mild to minimal depression bracket (PHQ-9). Consistent with these findings, a number of recent studies have shown improvements in affective symptoms following prebiotic interventions^(21-26)^, often accompanied by increases in *Bifidobacterium*^(21, 23, 26)^. In this study, we too observed significant increases in *Bifidobacterium* and *Parabacteroides* in the treatment group, both of which have been linked to fibre degradation and the subsequent production of SCFAs^(54, 55)^. It is important to note that we did not observe any between-group differences in these genera at week 12. This may in part be attributed to the larger sample size in the treatment group compared to control, which increased our power to detect within-group differences. SCFA production is thought to be one of the main mechanisms driving the relationship between prebiotic fibre intake and improvements in brain activity and affect. As part of this mechanism, SCFAs downregulate systemic inflammation by maintaining intestinal barrier integrity^(56, 57)^, promoting mucous production^(58)^, and regulating the secretion of interleukins^(17, 59)^. This is particularly relevant here, as pro-inflammatory cytokines can influence neuroinflammation and associated changes in brain activity and behaviour by crossing the BBB or interacting with the BBB interface^(7, 13)^. SCFAs also signal to the brain via enteroendocrine-mediated vagal signalling, offering a fast, direct, and accessible route to influence mood, stress, and anxiety^(18, 19)^. As a complementary mechanism, SCFAs have been shown to attenuate the stress response acting via the hypothalamic-pituitary-adrenal (HPA) axis and may modulate the relationship between prebiotic fibre and perceived stress^(60, 61)^.

Providing preliminary support for the above, our work suggests that changes in systemic inflammation may be closely associated with changes in mood and affective scores. Specifically, results from our stepwise multiple regression analyses showed that changes in CRP and GI symptoms could explain variability across all measures of depression, anxiety, and stress. The coefficient estimate for CRP was strongest in depression, where changes in CRP and GSRS (total) explained 35% of the variability in DASS-D scores. The strength of this effect is consistent with previous work assessing inflammatory cytokines with depressive symptoms^(62, 63)^, and functional connectivity changes in depression-related brain networks^(64, 65)^. More broadly, this finding is in line with the cytokine-depression response, where acute inflammatory responses can manifest as social avoidance, anhedonia, fatigue, and depressed mood^(8-10)^. The contribution of GI symptoms (GSRS) to depression, anxiety, and stress scores were also a consistent feature across regression models. GI symptoms have been recognised as a risk factor for poor mental health. That is, while constipation is generally considered a sequalae of depression or a side-effect from anti-depressant medication, a recent study shows it may be an independent risk factor or a prodromal symptom of depression^(66)^. Taken together, our results suggest two overlapping routes via which peripherally related signals may influence mood and affect: by regulating inflammation, and/or directly and indirectly improving GI symptoms.

The strengths of our investigation include the RCT design, high tolerability and adherence to the intervention, and no SAEs reported. However, several caveats need to be considered. As a pilot study, our sample size is relatively small and limited by the unbalanced randomisation allocation. While this provided necessary insights about the tolerability and side-effects from using the prebiotic blend, a future extension of this work should involve a larger, placebo-controlled, and balanced study design. Another consideration is that our control group did not take a placebo, which limits us from identifying placebo effects from taking the prebiotic blend. However, this is the first study to show multivariate associations between biologically relevant markers, including the gut microbiome and CRP, with depression, stress, and anxiety. This has enabled us to see considerable consistency with previous observational and interventional studies. Also, although the treatment and control groups did not statistically differ in baseline characteristics, the control group reported more GI and depression scores, hence, our findings should be interpreted with caution. While it is highly plausible that the production of SCFAs influenced the observed changes in primary and secondary outcomes, a further limitation in this study is that these metabolites were not quantified due to the de-centralised nature of this RCT. SCFAs are continually produced in and absorbed by the colonic epithelium; thus, future studies would benefit from measuring circulating SCFAs as they provide a better indication of prebiotic fermentation than faecal SCFAs^(60, 67)^.

## Conclusion

Underlying systemic inflammation has been shown to contribute to alterations in brain activity, behaviour, and affect. As such, there is a growing interest to develop novel, simple, and cost-effective interventions that can target systemic inflammation to improve long-term psychological health. Here, we showed that a 12-week long daily prebiotic fibre intervention lowered inflammation, modulated gut microbiome composition, and reduced sub-clinical levels of stress, anxiety, and depression in MetS. Larger and placebo controlled RCTs (controlling for various participant and sample characteristics)^(68)^ will be necessary to confirm these findings and explore the mechanisms that impact the gut microbiota, inflammation (ideally by using wide range of inflammation biomarkers measured at multiple time points to control for intra-individual variations), and the brain.

## Author contributions

Conceptualization and methodology, C.V. Hall, P. Hepsomali, B. Dalile, T. Gurry; Formal analysis: C.V. Hal and T.Gurry l; Data curation: C.V. Hall; Writing—original draft, review, and editing, all authors; Funding acquisition, T. Gurry; Supervision, T. Gurry and L. Scapozza.

## Data Availability

All data produced in the present study are available upon reasonable request to the authors.

