## Supplementary Material for "Effects of a diverse prebiotic fibre blend on inflammation, the gut microbiota, and affective symptoms: A pilot open label randomised controlled trial"

**Hall et al.**

**Supplementary Note 1.** Detailed eligibility criteria

Individuals aged 18-75 years with a diagnosis of Metabolic Syndrome (MetS), yet not receiving treatment for their symptoms, were recruited across the UK. MetS was diagnosed using the IDF criteria (1). MetS is defined as having abdominal obesity (waist circumference ≥ 94 cm in men, and ≥ 80 cm in women) plus two or more of the following: raised triglycerides (≥1.7 mmol/L); reduced HDL-C (< 1.03 mmol/L in men and < 1.29 mmol/L in women); raised systolic blood pressure (≥ 130 mmHg); raised diastolic blood pressure (≥ 85 mmHg); treatment of previously diagnosed hypertension; raised fasting plasma glucose (≥ 5.6 mmol/L); or previously diagnosed with pre-diabetes. Additional inclusion criteria included body mass index < 45 kg/m2, capacity to give informed consent, and ability to comply with study requirements. Exclusion criteria included a current diagnosis of Type 1 or 2 diabetes or cardiovascular disease; receiving medications that lower cholesterol, blood pressure, or blood glucose levels; pregnancy, lactation, or an intent to become pregnant during the course of the study; continuous antibiotic use for > 3 days within 1 month prior to enrolment; continuous use of weight-loss drug for > 1 month before screening; major change in dietary intake in past month (e.g. excluding whole food groups); currently consuming large doses of prebiotic or probiotic supplements; prior use (< 6 months) of any blood glucose or cholesterol lowering medication; a significant gastrointestinal (GI) condition affecting absorption including (but not limited to) inflammatory bowel disease; weight loss surgery; irritable bowel disease; end stage renal disease; active cancer, or treatment for any cancer, in last 3 years. Eligible participants were recruited through email and online campaigns. Participants were withdrawn from the trial if they met one of the exclusion criteria during the study e.g., starting a medication to control their symptoms.

**Supplementary Note 2.** Nutritional breakdown of the diverse prebiotic supplement

Each 10g daily serve of supplement contains the following fibres:

- Fructooligosaccharides
- Inulin
- Resistant dextrin
- Resistant maltodextrin
- Partially hydrolysed guar gum
- Guar gum

**Supplementary Note 3.** Example of GP healthy eating guidelines

The dietary and exercise recommendations were provided to participants via an online GP dashboard. Recommendations were slightly personalised, but included a variation of the following:

- Consume a diversity of vegetables and fruit daily
- Eat at least 2 portions of fish per week, including a portion of oily fish. Oily fish like salmon, tuna and mackerel are a great source of Omega 3. Consider taking an Omega-3 supplement to improve your Omega 3 levels.
- If you’re on a plant-based diet, your major source of Omega 3s will be seeds such as chia seeds, flax seeds, unsalted nuts and leafy greens such as broccoli, spinach and kale.
- It is also important to limit your intake of Omega 6 fats. These are found in sunflower and vegetable oils, as well as processed foods and refined grains.
- Reduce intake of sugar and food products containing refined sugars including fructose. This is more important to avoid than fat.
- Avoid white starchy foods such as white bread, pasta.
- Replace potatoes for sweet potatoes.
- Choose to consume wholegrain varieties of starchy food like oats.
- Avoid processed foods such as pies, biscuits and fried food as these contain harmful trans fats.
- Eat at least 4 to 5 portions of unsalted nuts, seeds, and legumes (such as chickpeas or beans) per week.

They were also provided with the UK Heart Healthy Guidelines, found here:

<https://www.heartuk.org.uk/downloads/health-professionals/publications/healthy-eating-guide.pdf>

**Supplementary Table 1**. Nutritional composition of fibre intervention

| **Typical Values** | **Per 100g** | **Per portion (10g)** | **% DV** |
| --- | --- | --- | --- |
| Energy | 185 kcal  774 kJ | 20 kcal  85 kJ | - |
| Fat | 0.0g | 0.0g | 0% |
| Carbohydrate  From sugar  Dietary fibre | 100g  1.4g  98.6g | 10g  0.1g  9.8g | 3%  0.4%  35% |
| Protein | 0.0g | 0.0g | 0% |
| Salt | 0.0g | 0.0g | 0% |

**Supplementary Table 2.** Results from multiple regression sensitivity analyses

| **Model** | **F-statistic** | **R^2^** | **Adjusted R^2^** | ***p*-values** |
| --- | --- | --- | --- | --- |
| PSS | 7.28 | 0.25 | 0.22 | **4.23 x 10^-5^** |
| DASS-A | 20.0 | 0.41 | 0.39 | **5.99 x 10^-10^** |
| DASS-D | 14.19 | 0.40 | 0.37 | **6.21 x 10^-9^** |
| DASS-S | 6.92 | 0.24 | 0.21 | **7.211 x 10^-5^** |

Perceived Stress Scale (PSS); Depression, Anxiety, and Stress Scale 42-item (DASS)-Stress (DASS-S); DASS-Anxiety (DASS-A); DASS-Depression (DASS-D). All p-values are Bonferroni-corrected.

**Supplementary Table 3.** Baseline, Week 12, and Delta (Week 12 – Baseline) results for clinical and affective outcome

|  | **Treatment** | | | **Control** | | |  |
| --- | --- | --- | --- | --- | --- | --- | --- |
|  | Baseline | Week 12 | Delta | Baseline | Week 12 | Delta | p-value |
| CRP (mmol/L) | 1.41 (0-14.8) | 1.82 (0.15-13.3) | -0.58 (-9.96-2.63) | 1.28 (0.15-30.6) | 2.36 (0.15-32.6) | 0.37 (-3.64-3.32) | **0.03** ^a^ |
| PSS | 17.85 ± 6.67 | 13.34 ± 5.83 | -4.43 ± 4.76 | 19.36 ± 5.68 | 19.00 ± 5.26 | -0.54 ± 3.45 | **0.004** ^b^ |
| DASS-D | 7.28 ± 9.34 | 4.00 ± 4.49 | -3.30 ± 8.04 | 13.21 ± 13.89 | 11.14 ± 13.13 | 0.08 ± 3.37 | **0.05** ^b^ |
| DASS-A | 5.25 ± 5.71 | 2.71 ± 3.47 | -2.32 ± 4.60 | 8.64 ± 9.68 | 7.43 ± 9.22 | 0.00 ± 2.30 | **0.03** ^b^ |
| DASS-S | 12.28 ± 9.96 | 7.08 ± 7.81 | -5.14 ± 7.09 | 13.43 ± 11.83 | 11.36 ± 10.80 | -0.17 ± 4.61 | **0.009** ^b^ |
| GAD-7 | 6.72 ± 5.39 | 3.87 ± 3.49 | -2.70 ± 3.93 | 8.29 ± 5.55 | 8.13 ± 5.44 | -0.15 ± 3.29 | **0.03** ^b^ |
| PHQ-9 | 7.50 ± 6.14 | 4.55 ± 3.90 | -2.92 ± 4.86 | 9.29 ± 7.02 | 8.20 ± 5.71 | -1.23 ± 4.71 | 0.28 ^b^ |
| GSRS-Total | 1.85 ± 0.58 | 1.54 ± 0.44 | -0.34 ± 0.61 | 2.33 ± 1.14 | 2.21 ± 1.02 | -0.06 ± 0.41 | 0.08 ^b^ |
| GSRS-Reflux | 1.42 ± 0.83 | 1.36 ± 0.77 | -0.09 ± 0.83 | 1.96 ± 1.54 | 1.93 ± 1.27 | 0.00 ± 1.04 | 0.77 ^b^ |
| GSRS-Indigestion | 2.38 ± 0.92 | 2.02 ± 0.91 | -0.41 ± 1.09 | 2.93 ± 1.79 | 2.57 ± 1.38 | 0.16 ± 0.65 | 0.70 ^b^ |
| GSRS-Pain | 1.70 ± 0.63 | 1.32 ± 0.43 | -0.41 ± 0.63 | 1.76 ± 0.73 | 1.89 ± 0.77 | 0.23 ± 0.53 | **0.002** ^b^ |
| GSRS-Constipation | 2.17 ± 1.59 | 1.67 ± 0.76 | -0.52 ± 1.67 | 2.64 ± 1.22 | 2.42 ± 1.10 | -0.15 ± 1.34 | 0.43 ^b^ |
| GSRS-Diarrhoea | 1.56 ± 0.72 | 1.32 ± 0.51 | -0.26 ± 0.63 | 2.33 ± 1.88 | 2.22 ± 1.73 | -0.05 ± 0.64 | 0.32 ^b^ |
| Systolic BP (mmHg) | 133 ± 14.8 | 126 ± 17.5 | -7.19 ± 19.4 | 128 ± 11.4 | 130 ± 14.0 | 2.23 ± 10.1 | **0.03** ^b^ |
| Diastolic BP (mmHg) | 84.0 ± 7.5 | 83.3 ± 12.5 | -0.28 ± 13.1 | 81.1 ± 10.5 | 82.8 ± 8.30 | 0.46 ± 6.13 | 0.79 ^b^ |
| Cholesterol (total) (mmol/L) | 5.95 ± 0.96 | 5.81 ± 1.16 | -0.05 ± 0.62 | 5.48 ± 1.27 | 5.50 ± 0.97 | -0.18 ± 1.17 | 0.72 ^b^ |
| Triglycerides (mmol/L) | 2.00 ± 1.19 | 1.82 ± 0.84 | -0.07 ± 0.50 | 1.94 ± 1.14 | 1.61 ± 0.72 | -0.40 ± 0.62 | 0.11 ^b^ |
| LDL-C (mmol/L) | 3.71 ± 0.90 | 3.63 ± 0.97 | -0.01 ± 0.60 | 3.34 ± 1.28 | 3.38 ± 0.85 | -0.15 ± 1.10 | 0.67 ^b^ |
| HDL-C (mmol/L) | 1.43 ± 0.41 | 1.36 ± 0.34 | -0.09 ± 0.21 | 1.44 ± 0.42 | 1.38 ± 0.46 | -0.06 ± 1.81 | 0.62 ^b^ |

Mean ± SD; Median (range); ^a^ Wilcoxon signed-rank test; ^b^ Unpaired t-test. C-reactive protein (CRP); Perceived Stress Scale (PSS); Depression, Anxiety, and Stress – 42-item scale (DASS) - Depression (DASS-D); DASS-anxiety (DASS-A); DASS-stress (DASS-S); Gastrointestinal Symptom Rating Scale (GSRS); blood pressure (BP); low-density lipoprotein - cholesterol (LDL-C); high-density lipoprotein - cholesterol (HDL-C).

**Supplementary Table 4.** Adverse events reported in the treatment group

| **Type of Adverse Event** | ***n*** |
| --- | --- |
| Wind | 19 |
| Bloating | 9 |
| Mild constipation | 7 |
| Abdominal pain | 6 |
| Mild diarrhoea | 5 |
| Burping | 1 |
